# Clinical course and risk factors for mortality of COVID-19 patients with pre-existing cirrhosis: A multicenter cohort study

**DOI:** 10.1101/2020.04.24.20072611

**Authors:** Xiaolong Qi, Yanna Liu, Jonathan A. Fallowfield, Jitao Wang, Jianwen Wang, Xinyu Li, Jindong Shi, Hongqiu Pan, Shengqiang Zou, Hongguang Zhang, Zhenhuai Chen, Fujian Li, Yan Luo, Mei Mei, Huiling Liu, Zhengyan Wang, Jinlin Li, Hua Yang, Huihua Xiang, Xiaodan Li, Tao Liu, Ming-Hua Zheng, Chuan Liu, Yifei Huang, Dan Xu, Xiaoguo Li, Ning Kang, Qing He, Ye Gu, Guo Zhang, Chuxiao Shao, Dengxiang Liu, Lin Zhang, Xun Li, Norifumi Kawada, Zicheng Jiang, Fengmei Wang, Bin Xiong, Tetsuo Takehara, Don C. Rockey, for the COVID-Cirrhosis-CHESS Group

**Author notes:** **Correspondence to:** Xiaolong Qi, M.D., Professor of Medicine, Chairman, CHESS (Chinese Portal Hypertension Diagnosis and Monitoring Study Group) Chief, CHESS Center, Institute of Portal Hypertension, The First Hospital of Lanzhou University, Lanzhou 730000, China, OR Bin Xiong, M.D., Professor of Medicine, Department of Radiology, Union Hospital, Tongji Medical College, Huazhong University of Science and Technology, Wuhan, China. Joint first authors (Xiaolong Qi, Yanna Liu, Jonathan A. Fallowfield, Jitao Wang). Joint senior authors (Xiaolong Qi, Tetsuo Takehara, Don C. Rockey). **Author contributions:** Concept and design: Xiaolong Qi; Acquisition and interpretation of data: Bin Xiong, Jianwen Wang, Xinyu Li, Jindong Shi, Hongqiu Pan, Shengqiang Zou, Hongguang Zhang, Zhenhuai Chen, Fujian Li, Yan Luo, Mei Mei, Huiling Liu, Zhengyan Wang, Jinlin Li, Hua Yang, Huihua Xiang, Xiaodan Li, Tao Liu, Ming-Hua Zheng, Chuan Liu, Yifei Huang, Dan Xu, Qing He, Ye Gu, Guo Zhang, Chuxiao Shao, Dengxiang Liu, Lin Zhang, Xun Li, Zicheng Jiang, Fengmei Wang; Drafting of the manuscript: Yanna Liu, Jitao Wang, Xiaolong Qi. Critical revision of the manuscript: Don C. Rockey, Jonathan A. Fallowfield, Tetsuo Takehara, Norifumi Kawada. **Conflicts of interest:** The authors have declared no conflict of interest related to the study. **Financial support:** None.

## Abstract

**Background:** Patients with pre-existing cirrhosis are considered at increased risk of severe coronavirus disease 2019 (COVID-19) but the clinical course in these patients has not yet been reported. This study aimed to provide a detailed report of the clinical characteristics and outcomes among COVID-19 patients with pre-existing cirrhosis.

**Methods:** In this retrospective, multicenter cohort study, we consecutively included all adult inpatients with laboratory-confirmed COVID-19 and pre-existing cirrhosis that had been discharged or had died by 24 March 2020 from 16 designated hospitals in China.

Demographic, clinical, laboratory and radiographic findings on admission, treatment, complications during hospitalization and clinical outcomes were collected and compared between survivors and non-survivors.

**Findings:** Twenty-one patients were included consecutively in this study, of whom 16 were cured and 5 died in hospital. Seventeen patients had compensated cirrhosis and hepatitis B virus infection was the most common etiology. Lymphocyte and platelet counts were lower, and direct bilirubin levels were higher in patients who died than those who survived (p= 0·040, 0·032, and 0·006, respectively). Acute respiratory distress syndrome and secondary infection were both the most frequently observed complications. Only one patient developed acute on chronic liver failure. Of the 5 non-survivors, all patients developed acute respiratory distress syndrome and 2 patients progressed to multiple organ dysfunction syndrome.

**Interpretation:** Lower lymphocyte and platelet counts, and higher direct bilirubin level might represent poor prognostic indicators in SARS-CoV-2-infected patients with pre-existing cirrhosis.

## Introduction

The coronavirus disease 2019 (COVID-19), caused by severe acute respiratory syndrome coronavirus-2 (SARS-CoV-2), has become a global challenge since the December 2019.^1-3^ Concerns have been raised recently about patients with pre-existing cirrhosis, who are known to have compromised immune function and poorer outcomes from acute respiratory distress syndrome (ARDS) than patients without cirrhosis.^1-3^ However, it remains unclear how SARS-CoV-2 infection affects the liver, if it modifies the clinical course of cirrhosis, and whether cirrhosis should be considered a significant risk factor for adverse outcomes of COVID-19.

In previous studies, the proportion of patients with pre-existing liver conditions (including hepatitis virus infection, abnormal liver function and chronic liver disease) among COVID-19 infected patients ranged from 2·0% to 11·0%.^4-8^ More than one half of patients admitted to hospital with SARS-CoV-2 infection may have elevated liver function tests and emerging liver function abnormalities after admission were associated with longer hospital stay.^9^ So far, no studies have reported the clinical features and risk factors for in-hospital death in COVID-19 patients with pre-existing cirrhosis. The aim of our multicenter study is to describe the demographic characteristics, coexisting conditions, laboratory and imaging findings, and outcomes among COVID-19 patients with pre-existing cirrhosis.

## Methods

### Study design and participants

This retrospective multicentre study (COVID-Cirrhosis-CHESS, ClinicalTrials.gov, NCT04329559) included consecutive patients with laboratory-confirmed COVID-19 and pre-existing cirrhosis from 16 designated hospitals in China between 31 December 2019 and 24 March 2020. A confirmed case of COVID-19 was defined by a positive result on a high-throughput sequencing or real-time reverse-transcriptase polymerase-chain-reaction assay from nasopharyngeal swab specimens; or a SARS-CoV-2 specific IgM and IgG in serum; or a SARS-CoV-2 specific IgG titration of at least 4-fold higher in the recovery stage than in the acute stage.^4,5,10^ Cirrhosis was diagnosed based on previous liver biopsy results and/or clinical findings, including a history of chronic liver disease as well as documented complications of chronic liver disease (ie, ascites, varices, hepatic encephalopathy) and/or imaging consistent with cirrhosis.^11^ Pregnant women and children (age < 18 years) were excluded. The study was approved by the Institutional Ethics Commission of The First Hospital of Lanzhou University (No. LDYYLL2020-37). Informed consent was waived, and researchers analyzed only deidentified data.

### Data collection

Demographic data, comorbidities, history of cirrhosis, onset symptoms or signs of COVID-19, laboratory findings and chest CT results on admission were collected. Child-Pugh class, and model for end-stage liver disease (MELD) score were assessed based on laboratory findings on admission. Treatment regimens, complications and clinical outcomes during hospitalization were also collected. According to the clinical outcomes, patients were divided into two groups: a survivor group and a non-survivor group.

### Study definition

Exposure history was defined as exposure to people with confirmed SARS-CoV-2 infection or to Wuhan City in China.^12^ Leukopenia was defined as a leukocyte count of less than 4×10^9^/L. Lymphocytopenia was defined as a lymphocyte count of less than 1×10^9^/L. Thrombocytopenia was defined as a platelet count of less than 100×10^9^/L. Hypoproteinemia was defined as a serum albumin level less than 35g/L. Hyperbilirubinemia was defined as a total bilirubin level higher than 17·1 μmol/L. The upper limit of the normal range of prothrombin time, aspartate transaminase, alanine aminotransferase, and gamma-glutamyl transferase were 12·5 seconds, 40U/L, 40U/L, and 50U/L, respectively.^13^ Secondary infection was diagnosed based on clinical symptoms or signs of nosocomial pneumonia, with a positive result of a new bacterial or fungal pathogen from a lower respiratory tract specimen or blood samples taken ≥48 h after admission. Ascites was defined as a case that had newly occurring or worsening ascites during hospitalization. Upper gastrointestinal bleeding was defined as patients who had overt signs of acute upper gastrointestinal bleeding (hematemesis, melena, or both).^14^ Septic shock was defined according to the World Health Organization criteria for COVID-19.^15^ Acute kidney injury and was defined according to the clinical guideline by the Kidney Disease: Improving Global Outcomes acute kidney injury work group.^16^

Acute-on-chronic liver failure (ACLF) was defined according to the European Association for the Study of the Liver criteria.^17^ ARDS was diagnosed according to the Berlin Definition.^18^

### Statistical analysis

Categorical variables were reported as counts and percentages. Continuous variables were presented as median and interquartile range. Mann-Whitney U test or Fisher’s exact test were used to compare differences between survivors and non-survivors where appropriate. A two-sided p-value of less than 0·05 was considered statistically significant. The data were analyzed by SPSS version 20·0 for Windows (SPSS Inc., Chicago, IL, USA).

## Results

Twenty-one COVID-19 patients with pre-existing cirrhosis were identified and all were included in the final analysis. The median age of the 21 patients was 68 years (range: 43-87 years) and 11 of them were male (Table 1). Most patients had compensated cirrhosis (81·0%) with Child-Pugh class A, B, and C in 16, 3, and 2 patients, respectively. The median MELD score was 8 (range: 6-23). Twelve patients had chronic hepatitis B, including 3 who also had hepatitis C virus co-infection, alcohol abuse and schistosomiasis, respectively. Comorbidities other than cirrhosis were present in most patients (66·7%), with hypertension (19·0%) and coronary heart disease (19·0%) being the most common comorbidities, followed by type 2 diabetes (14·3%) and malignancy (14·3%). The median interval between onset of the illness and admission was 8 days, ranging from 0 to 30 days. The most common symptoms on admission were fever and cough, followed by shortness of breath and sputum production. There were no significant differences between survivors (n=16) and non-survivors (n=5) on age, sex, comorbidities, etiology of cirrhosis, stage of cirrhosis (compensated versus decompensated), Child-Pugh class, MELD score, interval between onset and admission, or onset symptoms of COVID-19.

**Table 1.**
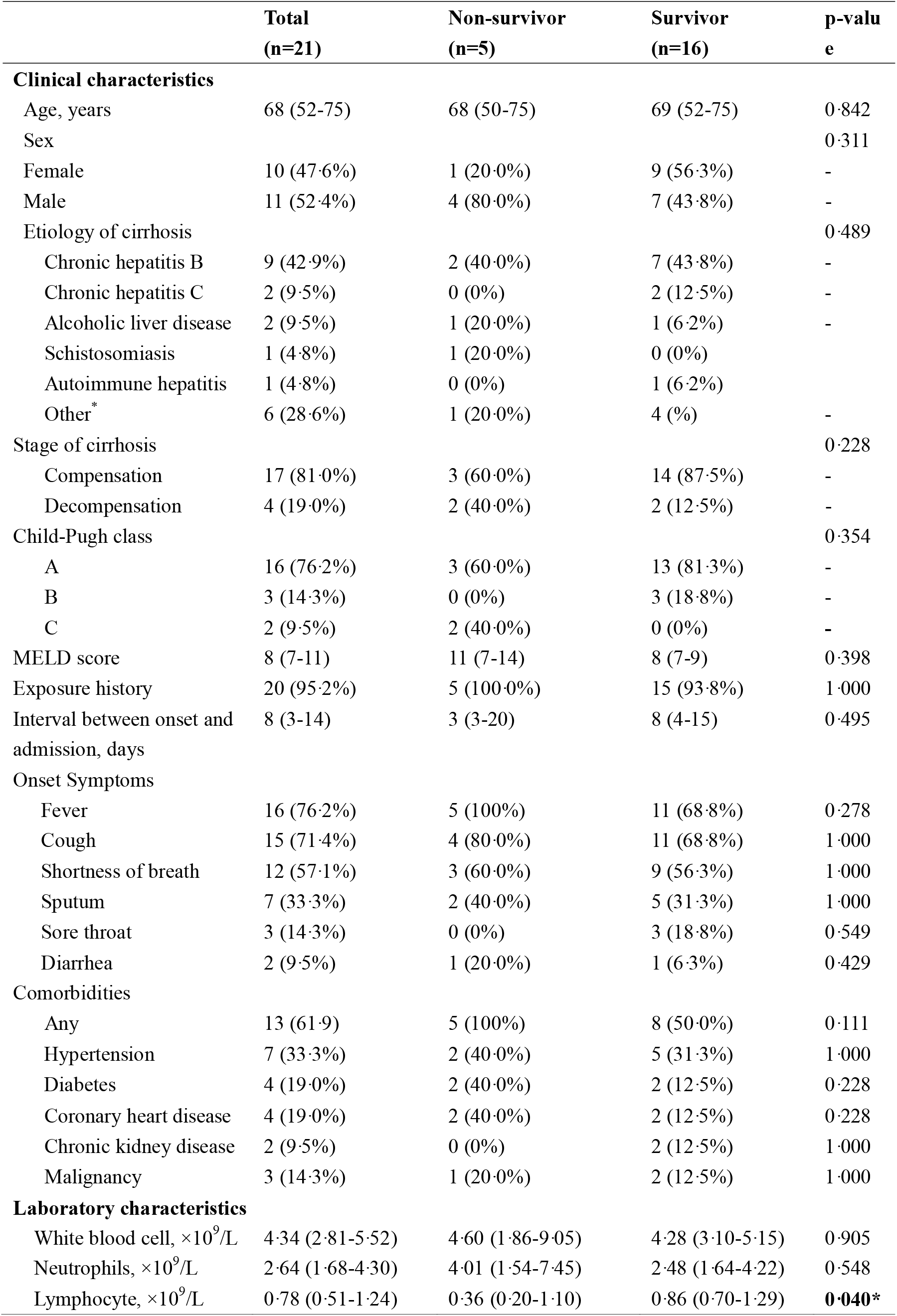
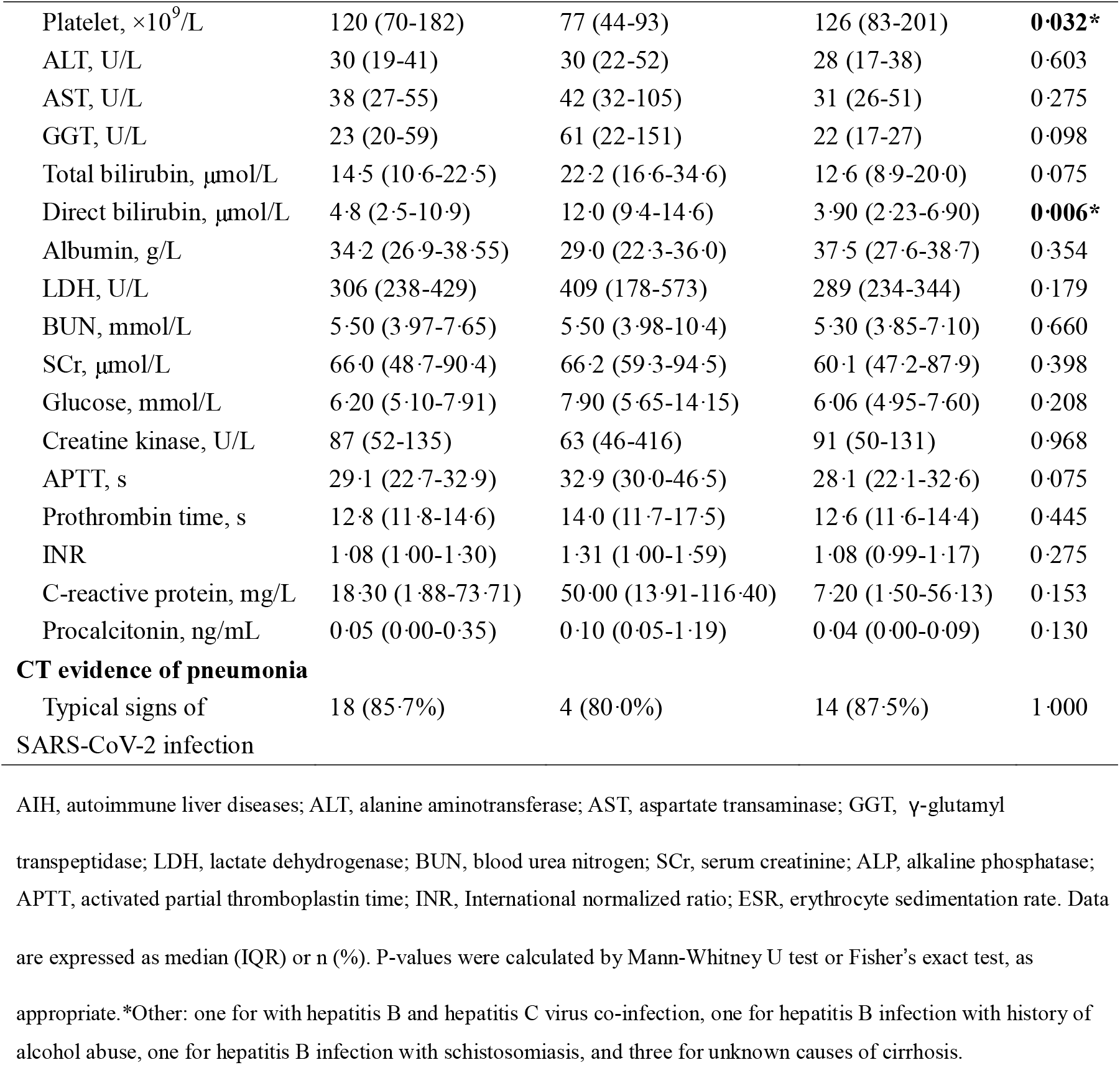
Clinical, laboratory, and radiographic findings on admission.

Leukopenia, lymphopenia and thrombocytopenia occurred in 8 (38·3%), 15 (71·4%), and 8 (38·1%) patients, respectively (Table 1). Hypoproteinemia, hyperbilirubinemia, and prolonged prothrombin time occurred in 11 (52·4%), 8 (38·1%), and 5 (23·8%) patients, respectively. Elevations in aspartate transaminase, alanine aminotransferase, and gamma-glutamyl transferase levels were present in 8 (38·1%), 5 (23·8%), and 5 (23·8%) patients, respectively. Nearly all patients had at least one typical sign of SARS-CoV-2 infection on chest CT including ground-glass opacities, consolidation, and bilateral pulmonary infiltration. Figure 1 showed representative chest CT images of COVID-19 and abdominal images of cirrhosis from survivor and non-survivor groups. Patients who died had lower total lymphocyte and platelet counts, and higher direct bilirubin levels than patients who survived (p= 0·040, 0·032, and 0·006, respectively) (Table 1).

**Figure 1.**
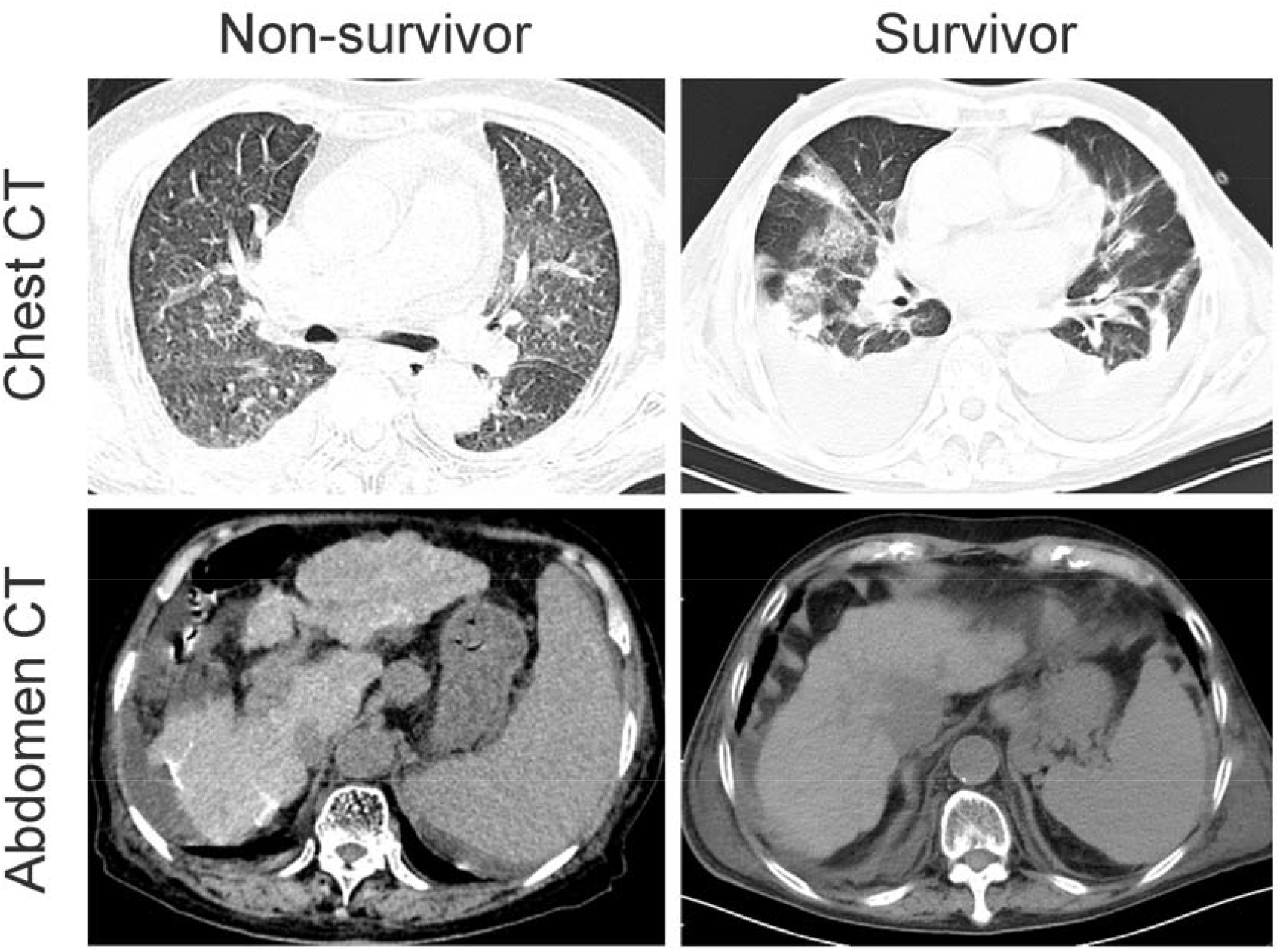
Baseline chest and abdomen CT scans of COVID-19 patients with pre-existing cirrhosis. Chest CT scans of both patients showed ground-glass opacities, consistent with SARS-CoV-2 infection. Abdominal CT scans showed abnormal liver morphology and splenomegaly, consistent with cirrhosis.

Treatment and complications occurring during hospitalization were summarized in Table 2. Five patients were transferred to the intensive care unit, including four patients from the non-survivor group (p= 0·004). Use of systemic corticosteroids (p= 0·003), non-invasive ventilation (p= 0·028), invasive mechanical ventilation (p= 0·008), continuous renal replacement therapy (p= 0·048), and extracorporeal membrane oxygenation (p= 0·048) differed significantly between non-survivors and survivors. Overall, 17 patients received antiviral treatment. Among them, arbidol monotherapy, lopinavir-ritonavir plus interferon alfa-2b combination therapy, and oseltamivir monotherapy were given to seven, five, and three patients, respectively. Two patients experienced a switch in the antiviral regimen (one from lopinavir-ritonavir plus arbidol combination therapy to interferon alfa-2b monotherapy; one from arbidol monotherapy to lopinavir-ritonavir plus interferon alfa-2b combination therapy). Secondary infection and ARDS were both the most frequently observed complication (28·6%), followed by ascites (23·8%). One patient developed ACLF. The frequency of ARDS and gastrointestinal bleeding were higher in non-survivors than survivors (100% vs. 6·3%, p< 0·001, and 60·0% vs. 6·3%, p= 0·028, respectively). Among the five patients who died, all developed ARDS and two of them progressed to multiple organ dysfunction syndromes.

**Table 2.**
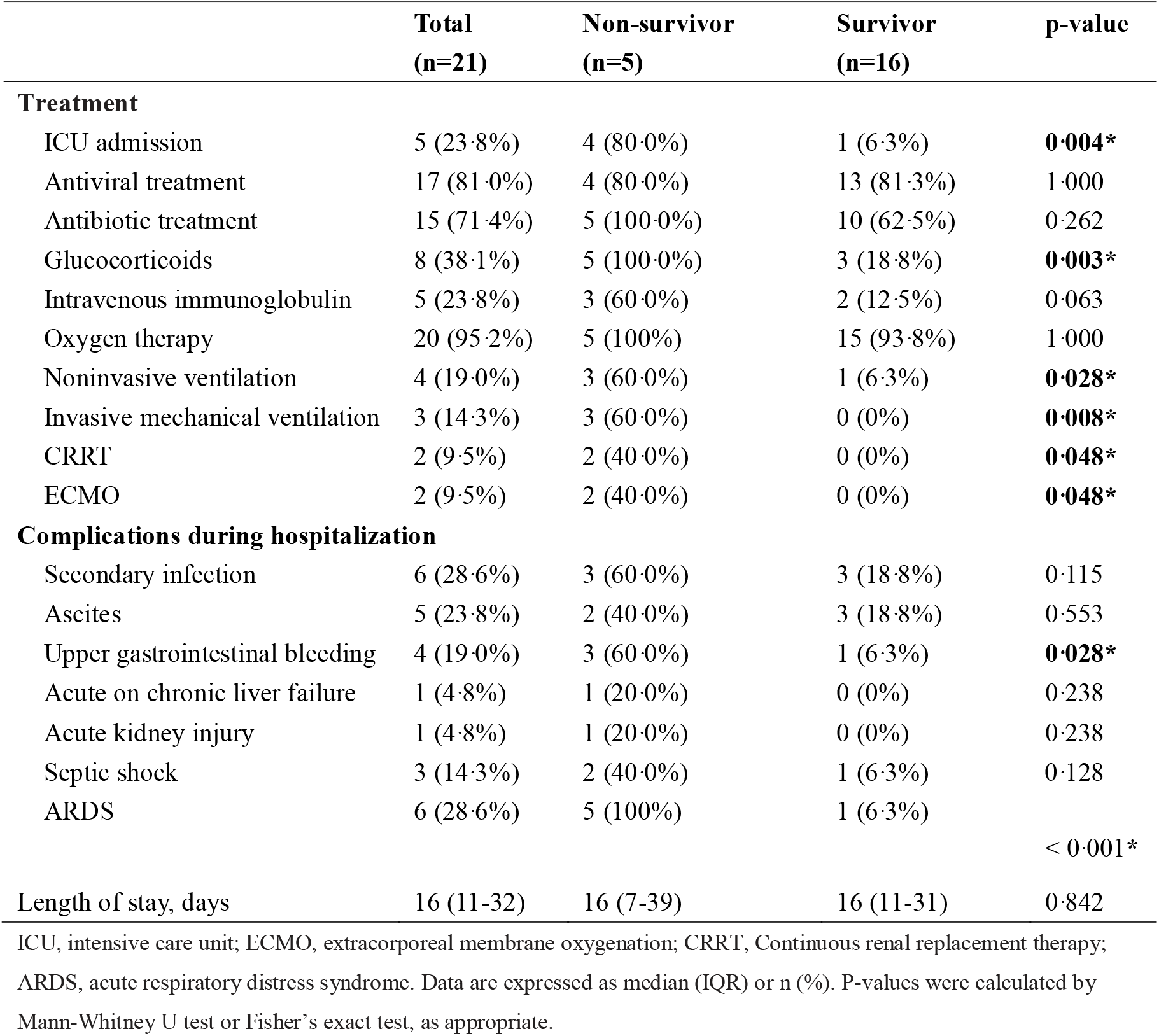
Treatment, complications, and outcomes.

## Discussion

This retrospective multicenter study (COVID-Cirrhosis-CHESS) reports the demographic characteristics, coexisting conditions, laboratory and imaging findings, and outcomes among COVID-19 patients with pre-existing cirrhosis. The cause of death in most patients is not due to progressive liver disease but rather pulmonary disease. Lower lymphocyte and platelet counts, and higher direct bilirubin level were observed in patients who survived compared to those who died.

In our cohort of COVID-19 patients with pre-existing cirrhosis, we found that fever and cough were the most common symptoms on admission; and most of the patients had comorbidities other than cirrhosis. These findings were similar to the previous studies of COVID-19 among general populations.^4-6^ According to the China Center for Disease Control and Prevention’s report of 44,672 laboratory-confirmed cases of COVID-19, deaths were mainly observed among patients of older age and/or with pre-existing comorbid conditions including cardiovascular disease, diabetes, chronic respiratory disease, hypertension, and cancer.^19^ In Italy, besides the above mentioned factors, male patients also had an increased risk of death, as revealed by the report of the first month of documented COVID-19 cases.^20^ So far, no studies have been reported risk factors for death in COVID-19 patients with pre-existing cirrhosis. Our analysis did not indicate a higher mortality in included patients of older age, male sex or with pre-existing comorbidities. This might be due to the narrow spectrum of our studied population and the limited sample size.

The liver plays important roles in both adaptive and innate immune responses as it is enriched with lymphocytes including T and B lymphocytes, and natural killer cells.^21^ In addition, the liver is a central regulator of the number of circulating platelets via thrombopoietin production and clearance of aged platelets.^22^ Thus, patients with cirrhosis are more likely to have lymphopenia and thrombocytopenia and, as the disease progresses, this becomes more prominent. In our study, the frequency of lymphopenia and thrombocytopenia were both as high as 80·0% in the non-survivor group, 40·0% of whom had decompensated cirrhosis. Additionally, a lower lymphocyte and platelet counts on admission were observed in non-survivors compared to survivors. These findings were also demonstrated in previous studies in the general population.^23,24^ A meta-analysis including 10 studies and 1,994 patients of COVID-19 showed that lymphopenia was the most common laboratory findings, which was consistent with respiratory virus infection.^23^ In another meta-analysis, studying the effect of platelet count on prognosis of COVID-19 showed that, comparing patients by survival, a low platelet count was associated with over five-fold increased risk of severe COVID-19.^24^ Direct bilirubin was also observed to be higher in non-survivors than survivors in our study, which suggests a potential adverse effect on liver function in this setting. These findings underscore that in patients with COVID-19 and pre-existing cirrhosis, a decrease in lymphocytes and platelet counts and increased direct bilirubin should be monitored as they may indicate a poorer prognosis.

We recognize limitations of the current study. First, the sample size is small, which limits analysis of predictive factors for mortality and the generalizability of the results. Further studies with large sample size and prospective design are needed to better describe the clinical course of COVID-19 in patients with pre-existing cirrhosis and to determine the risk factors for poor outcomes in this setting. Secondly, the majority of patients in this study had underlying hepatitis B virus-related cirrhosis. However, to date there is no evidence to suggest that patients with stable chronic liver disease due to viral hepatitis have increased susceptibility to SARS-CoV-2 infection. In the absence of a liver biopsy, we do not know whether SARS-CoV-2 infection had effects on liver histopathology in our study population. Lastly, the study included only patients from Chinese medical institutions, and whether the data are generalizable to patents in other parts of the world remains unknown.

In conclusion, we have reported the demographic characteristics, coexisting conditions, laboratory and imaging findings, and outcomes among COVID-19 patients with pre-existing cirrhosis. It appears that the cause of death in most patients is not due to progressive liver disease (i.e. development of ACLF), but rather pulmonary disease. At presentation, lower lymphocyte and platelet counts, and higher direct bilirubin level may be associated with higher risk of death.

## Data Availability

I confirm the availability of all data referred to in the manuscript

## Declaration of interests

We declare no competing interests related to this paper.

## Acknowledgement

We thank the great support and critical comments of Xavier Dray (Saint Antoine Hospital, APHP & Sorbonne University, France), Mingkai Chen (Renmin Hospital of Wuhan University, China), and Necati Örmeci (Ankara University School of Medicine, Turkey).

## Notes

### Competing Interest Statement

The authors have declared no competing interest.

### Clinical Trial

NCT04329559

